# Can we predict antibody responses in SARS-CoV-2? A cohort analysis

**DOI:** 10.1101/2021.03.15.21253267

**Authors:** Mary Gaeddert, Philip Kitchen, Tobias Broger, Stefan Weber, Ralf Bartenschlager, Anna Plaszczyca, Hans-Georg Kräusslich, Barbara Müller, Margarida Souto-Carneiro, Maike Janssen, Carsten Müller-Tidow, Uta Merle, Yannis Herrmann, Lukas Raedeker, Jakob Sebastian, Niall Brindl, Tim Starck, Claudia M. Denkinger

## Abstract

**Background:** After infection with severe acute respiratory syndrome coronavirus (SARS-CoV-2), Immunoglobulin G (IgG) antibodies and virus-specific neutralizing antibodies (nAbs) develop. This study describes antibody responses in a cohort of recovered COVID-19 patients to identify predictors.

**Methods:** We recruited patients with confirmed SARS-CoV-2 infection from Heidelberg, Germany. Blood samples were collected three weeks after COVID-19 symptoms ended. Participants with high antibody titers were invited for follow-up visits. IgG titers were measured by the Euroimmun Assay, and nAbs titers in a SARS-CoV-2 infection-based assay.

**Results:** 281 participants were enrolled between April and August 2020 with IgG testing, 145 (51.6%) had nAbs, and 35 (12.5%) had follow-up. The median IgG optical density (OD) ratio was 3.1 (Interquartile range (IQR) 1.6-5.1), and 24.1% (35/145) had a nAb titer>1:80. Higher IgG titers were associated with increased age and more severe disease, and higher nAbs were associated with male gender and CT-value of 25-30 on RT-PCR at diagnosis. The median IgG OD ratio on follow-up was 3.7 (IQR 2.9-5.9), a median increase of 0.5 (IQR −0.3-1.7). Six participants with follow-up nAbs all had titers ≤ 1:80.

**Conclusions:** While age and disease severity were correlated with IgG responses, predictive factors for nAbs in convalescent patients remain unclear.

## Introduction

An understanding of the robustness of the antibody response against severe acute respiratory syndrome coronavirus 2 (SARS-CoV-2) and its longevity is critical to determine the necessary levels and duration of antibody titers that confer protection against re-infection.

After infection with the severe acute respiratory syndrome coronavirus 2 (SARS-CoV-2), about two thirds of hospitalized patients show IgG seroconversion within the first two weeks of infection [1] while later it will be up to 90% [2]. In contrast for mild cases, seroconversion might only be observed in 48% [2]. Previous studies in hospitalized patients with Coronavirus disease 2019 (COVID-19) indicate some correlation between disease severity and strength of the antibody response [3-6]. Also, studies have reported varying results regarding the change of IgG antibodies over time, with some evidence that this time course may also be related to disease severity [6-8].

Virus-specific neutralizing antibodies (nAbs) have the ability to inhibit virus replication in an ongoing infection and are likely to play a role in preventing re-infection. It has been shown that SARS-CoV-2 infection can result in the production of antibodies with neutralizing properties in vitro and protective effects in animal models [9]. However, the precise correlation between IgG levels and nAb response [10], as well as the predictors of nAbs in an effective and durable immune response against SARS-CoV-2 [9, 11] have not been clarified. Similar to IgG levels, nAbs may decrease several months after infection, but tend to remain higher following symptomatic infection compared with asymptomatic [8].

Understanding the levels of IgG and nAbs that are relevant for protection is also important for the use of convalescent plasma in the treatment of SARS-CoV-2. Convalescent plasma has already been authorized by the FDA under Emergency Use Authorization [12, 13], however the impact on patient outcomes has been variable [14, 15]. There is therefore also an urgent need to identify factors that predict which convalescent patients will have robust immune responses to SARS-CoV-2 infection and be suitable to serve as plasma donors.

The aim of this study is to describe the antibody response in a cohort of recovered COVID-19 patients and identify predictors of total IgG and neutralizing antibody responses. The results will be useful in identifying the most suitable plasma donors and may further contribute to understanding the risk for future re-infection.

## Methods

### Study design and participants

Convalescent COVID-19 patients from the Rhein-Neckar region surrounding Heidelberg, Germany, were recruited for this study. Participants were identified by the local Public Health Authority as meeting the current guidelines for COVID testing (exposure to a SARS CoV-2 positive case, suggestive symptoms, or travel to a risk area) and were tested in a certified test center in the Heidelberg area. Those who tested positive for SARS-CoV-2 infection were contacted approximately two to three weeks after the positive test and asked to participate in the study. A survey conducted online or by telephone collected retrospective data on demographic and clinical information, including symptoms, hospitalization, and measures of COVID-19 disease severity. After survey completion, participants were screened for potential plasma donation. The inclusion criteria for plasma donation were age less than 65 years and no comorbidities. Participants from outside the Rhein Neckar region were also recruited through a public advertisement of the COVID-19 plasma donor program. These participants followed the procedures below but did not complete the survey and are missing the demographic and clinical information.

The first study visit was conducted at the Heidelberg University Hospital at least three weeks after symptoms ended. During this visit, a nasopharyngeal swab and blood samples were collected for testing as described below. Convalescent patients with high IgG levels (optical density (OD) ratio > 2.5) and a nAbs titer (> 1:80) were invited to participate as plasma donors. Serologic testing was performed again at the time of plasma donation.

All procedures were performed in accordance with the ethical standards of the institutional research committee of Heidelberg University (study approval number S686/2018). Written informed consent was obtained from each participant.

### Molecular Assays

Nasopharyngeal swabs (eSwabs, Copan) were analyzed with Allplex 2019-nCoV Assay (Seegene Inc, Seoul, South Korea) and Tib-Molbiol 2019-nCoV LightMix (TIB Molbiol, Berlin, Germany) for viral RNA using reverse-transcriptase polymerase chain reaction (RT-PCR) designed to detect RdRP and N genes specific for SARS-CoV-2 and the E gene for all of Sarbecovirus including SARS-CoV-2. Cycle threshold-values (CT-values) under 38 were considered positive.

### Serological Assay

Enzyme linked immunosorbent assay (ELISA) measurements for determination of IgG reactivity against the S1 domain of the viral spike (S) protein were carried out using the Euroimmun Anti-SARS-CoV-2-ELISA (IgG) (EI 2606-9601 G; EUROIMMUN Medizinische Labordiagnostika AG, Lübeck, Germany) test kit according to the manufacturer’s protocol. Samples were processed on an Euroimmun Analyzer I instrument according to the manufacturer’s instructions. Immunoreactivity was determined by measuring the optical density at 450 nm (OD450) and divided by the OD450 of a calibrator comprised in the kit to minimize inter-assay variation. The interpretation of the semi-quantitative ratiometric values obtained followed the manufacturer’s test instructions: ratios <0.8 were classified as negative, 0.8-1.1 as borderline, and ≥1.1 or higher as positive. Sera with an OD ratio >2.5 were tested for the presence of neutralizing antibodies. A subset of samples was also tested with an ELISA assay targeting viral nucleocapsid antigen using the EDI™ Novel Coronavirus COVID-19 IgG ELISA Kit (Epitope Diaganostics Inc, San Diego CA, USA), according to the manufacturer’s instructions. Samples were analyzed in the SpectraMax M2^e^ multi-detection microplate reader (Molecular Devices, USA) at 450nm. Samples with an O.D. at 450nm ≥0.22 were considered positive. To minimize inter-assay variation the O.D. values were divided by the kit’s internal calibration control. The interpretation of the semi-quantitative ratiometric values was carried out similarly as for the S1 domain of the viral spike protein.

### Neutralization assay

VeroE6 cells were seeded into 96-well plates one day before the assay. Serial dilutions of sera starting from a 1:10 dilution were prepared in OptiMEM (ThermoFisher Scientific, Waltham, MA, USA) in a final volume of 75 µl and incubated with ∼24,000 PFU of SARS-CoV-2 (BavPat1/2020 strain, European Virus Archive) per well for 1 hour at 37 °C in a final volume of 150 µl. One third of each serum-virus mixture (50 µl) was then used for infection (final MOI ∼0.25). Infections were performed in duplicates. At 20 hours post-infection, cells were fixed with 5% formaldehyde and immune-stained using a primary mouse antiserum binding to double-stranded RNA, a viral replication intermediate (J2 mouse antibody; Scicons, Szirák, Hungary) and a secondary anti-mouse horseradish peroxidase-coupled antibody (Merck, Darmstadt, Germany). The signal was developed using the KPL SureBlueTM TMB peroxidase substrate (Seracare, Milford, MA, USA) and measured in a plate reader at a wavelength of 450 nm. Data were normalized to no-serum control (100%) and mock-infected control (0%). The neutralization (NT) titer was defined as the highest serum dilution resulting in more than 50% reduction of the normalized signal.

### Statistical Analysis

We assessed factors associated with antibody response by analyzing demographic and clinical characteristics of participants with different levels of IgG and nAbs. Linear regression was performed to determine the association of demographic and clinical characteristics with IgG antibody titers as a continuous outcome. Logistic regression was performed to study the association of the same covariates with nAbs as a categorial outcome. For both regression analyses, factors with a p-value <0.1 in the univariate analysis were included in the multivariable model, and retained in the model if the p-value was <0.05 using manual backward elimination. IgG titers from convalescent and follow-up samples were compared using the Wilcoxon matched-pairs signed-ranks test. Analysis was performed using Stata version 15.1 (StataCorp LLC. College Station, TX).

### Definitions

The first swab collected by the Public Health Authority for initial diagnosis of SARS-CoV-2 infection is referred to as the ‘diagnostic swab’, and the second swab collected at the screening visit for plasma donation is referred to as the ‘convalescent swab’. An IgG OD ratio <1.1 was defined as negative, 1.1 to 2.5 as weak positive, and >2.5 as high positive. NAbs were categorized into high and low, with titers ≤ 1:80 defined as low and >1:80 as high by expert consensus after review of data from an inpatient cohort.

An illness severity score was created using a combination of self-reported activity restriction, the presence of COVID-19 symptoms indicative of severe disease (cough, fever, or dyspnea), and hospitalization. The categories were defined as mild illness (normal activity, none of the three symptoms, and not hospitalized), moderate illness (restricted activity, symptomatic, and not hospitalized), and severe illness (severely restricted activity, symptomatic, and hospitalized).

## Results

A total of 281 participants were enrolled between April and August 2020. Survey responses were available for 200 (71.2%) participants. The population was 55% (107/194) male with a median age of 39 ([Interquartile range] IQR 29-50) (Table 1). Only 3/171 (1.8%) participants were hospitalized, with one in intensive care. The median CT-value at the time of diagnosis was available for 165/281 and was 25 (IQR 20 to 28). IgG testing was performed for all 281 participants and of these, ODs were >2.5 in 145 (51.6%). Thirty-five participants (12.5%) had serology repeated at the time of plasma donation.

**Table 1.** **Demographic and clinical characteristic of participants with convalescent testing (n=281)**

|  | n/N | % |
| --- | --- | --- |
| **Demographics** |  |  |
| Age, median [IQR] | 39 [29-50] |  |
| Gender |  |  |
| Male | 107/194 | 55.2 |
| Female | 86/194 | 44.3 |
| Diverse | 1/194 | 0.5 |
| **Diagnosis** |  |  |
| CT value of diagnostic swab, median [IQR] | 25 [20-28] |  |
| CT value of diagnostic swab |  |  |
| <25 | 87/165<sup>a</sup> | 52.7 |
| 25-30 | 54/165 | 32.7 |
| >30 | 24/165 | 14.6 |
| **Convalescent testing** |  |  |
| Days from diagnosis to convalescent testing, median [IQR] | 34 [28-43] |  |
| Days from end of symptoms to |  |  |
| convalescent testing, median [IQR] | 29 [22-37] |  |
| Result of convalescent swab |  |  |
| Negative | 249/274 | 90.9 |
| Positive | 25/274 | 9.1 |
| CT value of convalescent swab if positive, median [IQR] | 37 [35-38] |  |
| IgG OD ratio, median [IQR] | 3.1 [1.6-5.1] |  |
| IgG result |  |  |
| Negative/Borderline (<1.1) | 43/281 | 15.3 |
| Weak positive (1.1-2.5) | 73/281 | 26.0 |
| High positive (>2.5) | 165/281 | 58.7 |
| Neutralizing Antibodies |  |  |
| Low ($\leq 1:80$) | 110/145 | 75.9 |
| High ($> 1:80$) | 35/145 | 24.1 |
| Neutralizing Antibodies if convalescent swab positive |  |  |
| Low ($\leq 1:80$) | 15/20 | 75.0 |
| High ($> 1:80$) | 5/20 | 25.0 |
| **Follow-up testing (N=35)** |  |  |
| Days from convalescent to follow-up blood draw, median [IQR] | 46 [23-57] |  |
| Follow-up IgG result, median [IQR] | 3.7 [2.9-5.9] |  |
| Follow-up IgG result |  |  |
| Negative/Borderline (<1.1) | 2/35 | 5.7 |
| Weak positive (1.1-2.5) | 5/35 | 14.3 |
| High positive (>2.5) | 28/35 | 80.0 |
| Change in IgG from convalescent to follow-up, median [IQR] | 0.5 [-0.3-1.7] |  |
| Follow-up Neutralizing Antibodies |  |  |
| Low ($\leq 1:80$) | 6/6 | 100.0 |

<sup>a</sup>Missing data: gender=87, CT value of diagnostic swab=116, result of convalescent swab=7, results of nAbs for convalescent swab-positive=5
\*participants missing results of the first swab were diagnosed outside of the University Hospital Heidelberg network)

Convalescent samples were collected from all participants a median of 34 days (IQR 28-43) after the diagnostic swab and 29 days (IQR 22 to 37) after the end of symptoms. The median IgG OD ratio at the convalescent visit was 3.1 (IQR 1.6 to 5.1), with 58.7% (165/281) of participants categorized as having a highly positive antibody response, 26.0% (73/281) with a positive response, and 15.3% 43/281 (15.3%) with a negative or borderline IgG antibody response. Only 24.1% of the tested participants (35/145) displayed a nAb titer >1:80. All three hospitalized participants displayed nAb titers of 1:160. The convalescent swab was positive by RT-PCR in 9.1% (25/274) of participants and the median CT value was 37 (IQR 35 to 38). Of those who had a positive convalescent swab, 25% (5/20) had nAbs >1:80.

The IgG convalescent titer was highest in adults 18 to 29 years old, decreasing in 30-to 44-year-olds, and increasing again in adults 45 to 60 years of age (Table 2 and Figure 1). The nAbs titers, in contrast, did not show a statistically significant change by age (Table 3 and Supplemental Figure 1). There was a correlation between increasing IgG titer and nAbs titer although Kappa was low (OR=1.6 ([95% Confidence Interval] 95%CI 1.3-1.9), p<0.001, Kappa=0.095) (Figure 2). The IgG OD ratio of convalescent sera was neither correlated with the CT-value of the diagnostic swab (p=0.666) (Supplemental Figure 2) nor with that of the convalescent swab (p=0.727).

**Table 2.** **Description of factors associated with Convalescent SARS CoV-2 IgG titers (n=200)**

| Characteristic<br>n/N (%) | Total<br>population | OD Ratio,<br>Median<br>[IQR] | Simple Linear<br>Regression<br>Coef (95%CI), p- | Multiple Linear<br>Regression<br>Coef (95%CI), p- |
| --- | --- | --- | --- | --- |
|  |  |  | value | value |
| **Demographic** |  |  |  |  |
| Age |  |  |  |  |
| 18-29 | 50/200 (25.0) | 3.7 [1.9-4.4] | 0.1 (-0.8;1.0), 0.829 | 0.2 (-0.7;1.0), 0.666 |
| 30-44 | 75/200 (37.5) | 2.6 [1.2-4.5] | Reference | Reference |
| 45-60 | 75/200 (37.5) | 3.6 [1.8-6.3] | 0.9 (0.2;1.7), **0.017** | 0.8 (0.05-1.6), **0.036** |
| Gender<sup>a</sup> |  |  |  |  |
| Male | 107/194 (55.2) | 3.1 [1.5-5.1] | 0.4 (-0.4;1.0), 0.405 |  |
| Female | 86/194 (44.3) | 3.3 [1.8-4.9] | Reference |  |
| Diverse<sup>b</sup> | 1/194 (0.5) | 8.4 [8.4-8.4] |  |  |
| **Clinical** |  |  |  |  |
| Activity Restriction |  |  |  |  |
| Normal activity | 51/188 (27.1) | 2.2 [1.2-3.9] | Reference | Reference |
| Minimal restriction | 81/188 (43.1) | 3.0 [1.8-4.3] | 0.5 (-0.3;1.4), 0.187 | 0.6 (-0.2;1.4), 0.163 |
| Moderate/Severe | 56/188 (29.8) | 5.0 [2.6-7.2] | 2.2 (1.3;3.1), **<0.001** | 2.1 (1.2;3.0), **<0.001** |
| Symptomatic |  |  |  |  |
| No | 4/188 (2.1) | 2.5 [0.9-4.0] | Reference |  |
| Yes | 184/188 (97.9) | 3.2 [1.8-5.1] | 1.2 (-1.2;3.7), 0.325 |  |
| Fever only |  |  |  |  |
| No | 118/192 (61.5) | 2.9 [1.5-4.5] | Reference |  |
| Yes | 74/192 (38.5) | 3.5 [2.1-5.9] | 0.9 (0.2;1.6), **0.010** |  |
| Fever, Dyspnea, or Cough |  |  |  |  |
| No | 76/192 (39.6) | 2.8 [1.6-4.4] | Reference |  |
| Yes | 116/192 (60.4) | 3.4 [1.9-5.6] | 0.7 (0.03;1.4), **0.040** |  |
| Symptom duration |  |  |  |  |
| 0-10 days | 60/200 (30.0) | 2.9 [1.6-4.5] | Reference |  |
| 11-20 days | 87/200 (43.5) | 3.5 [2.1-5.7] | 0.9 (0.1;1.7), **0.034** |  |
| >20 days | 53/200 (26.5) | 2.9 [1.4-5.0] | 0.2 (-0.7;1.1), 0.731 |  |
| **Convalescent testing** |  |  |  |  |
| Days from Symptom onset to convalescent testing |  |  |  |  |
| 0-35 | 63/180 (35.0) | 2.6 [1.5-4.9] | Reference |  |
| 35-50 | 89/180 (49.4) | 3.4 [2.0-5.5] | 0.6 (-0.2;1.4), 0.145 |  |
| >50 | 28/180 (15.6) | 3.5 [1.6-6.4] | 0.7 (-0.4;1.8), 0.182 |  |
| CT value of diagnostic swab |  |  |  |  |
| <25 | 79/151 (52.3) | 3.3 [2.1-4.9] | -0.1 (-1.2;1.0), 0.880 |  |
| 25-30 | 48/151 (31.8) | 3.6 [1.6-5.8] | 0.3 (-0.9;1.6), 0.569 |  |
| >30 | 24/151 (15.9) | 3.5 [1.9-4.8] | Reference |  |
| Result of convalescent swab |  |  |  |  |
| Negative | 177/199 (88.5) | 3.0 [1.6-5.1] | Reference |  |
| Positive | 22/199 (11.1) | 3.8 [2.2-4.7] | 0.2 (-0.9;1.2), 0.780 |  |

<sup>a</sup>Missing data: gender=6, Activity Restriction n=12; Symptomatic n=12; specific symptoms n=8; start of symptoms n=20; diagnostic swab n=49; convalescent swab n=1
<sup>b</sup>Diverse gender (n=1) excluded from regression model

**Table 3.**
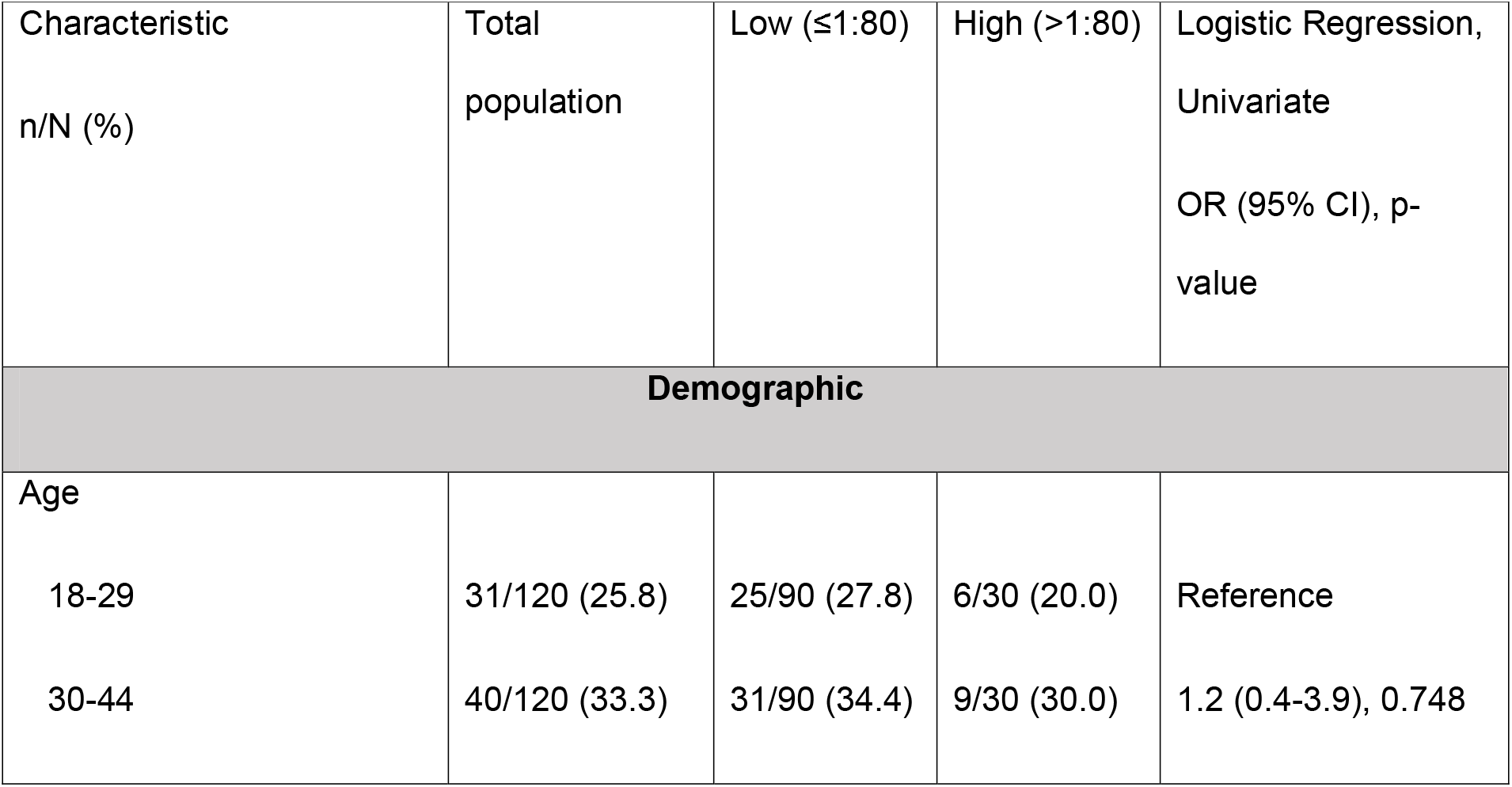
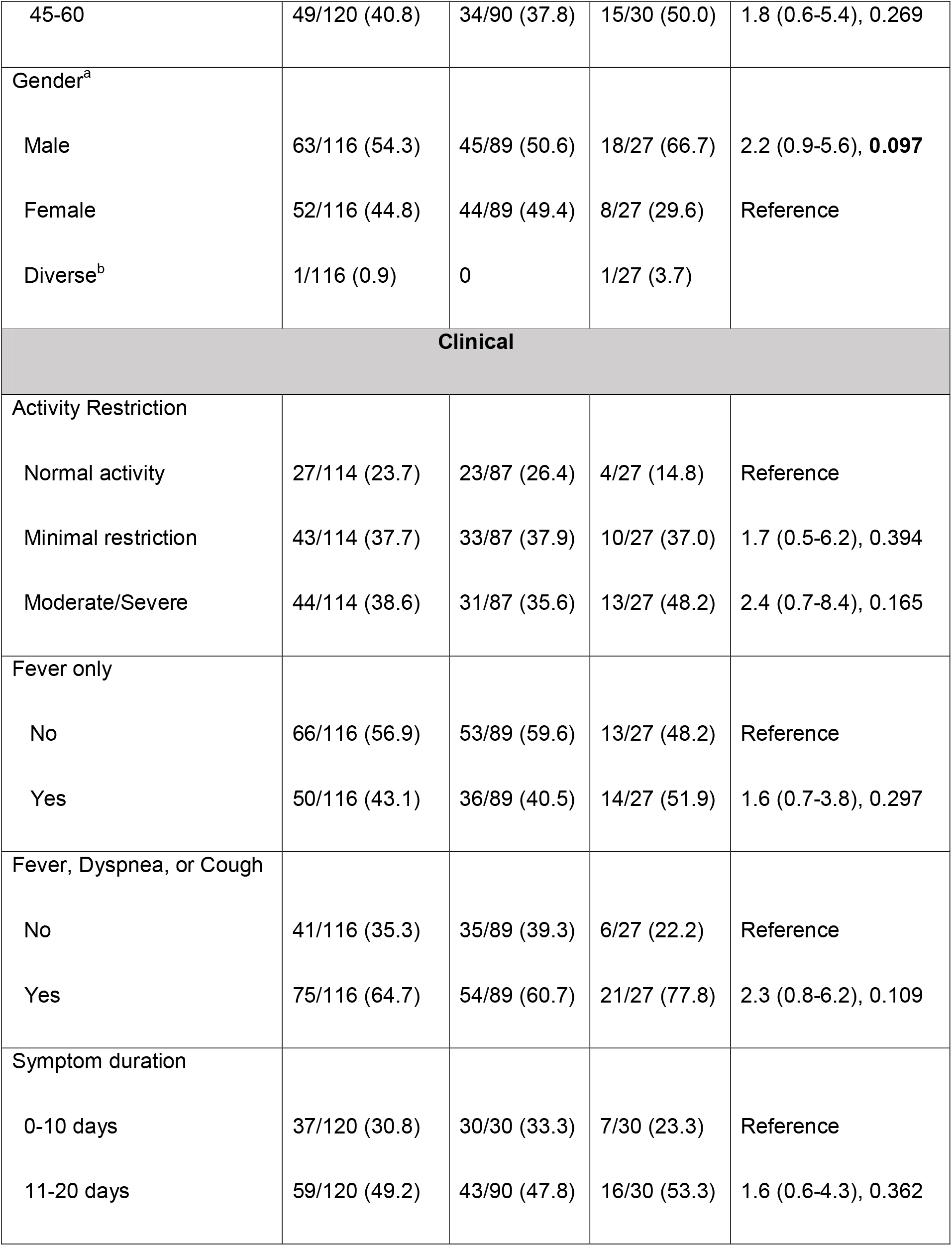
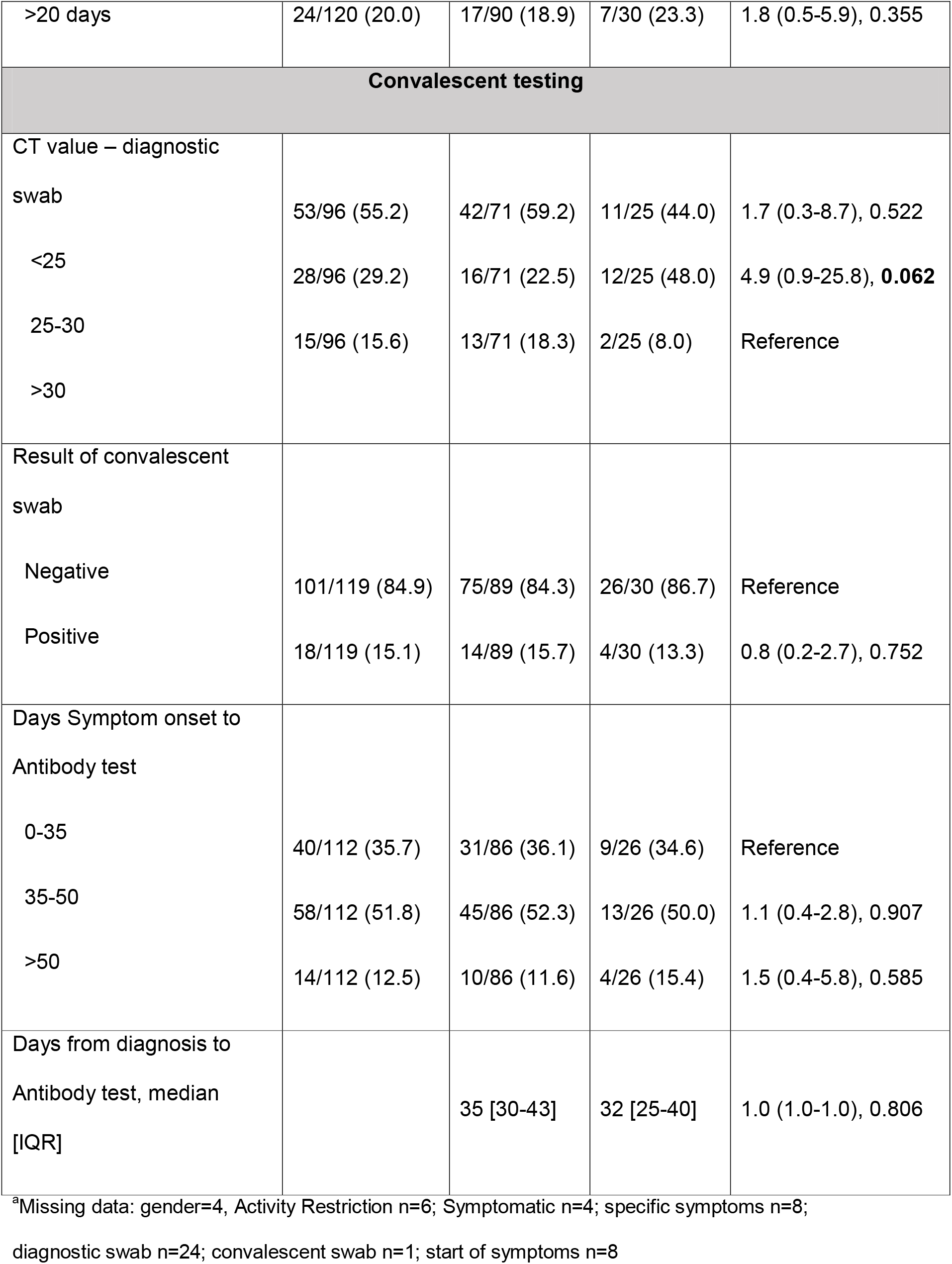
**Description of factors associated with Convalescent SARS CoV-2 Neutralizing antibody titers (n=120)**

**Figure 1.**
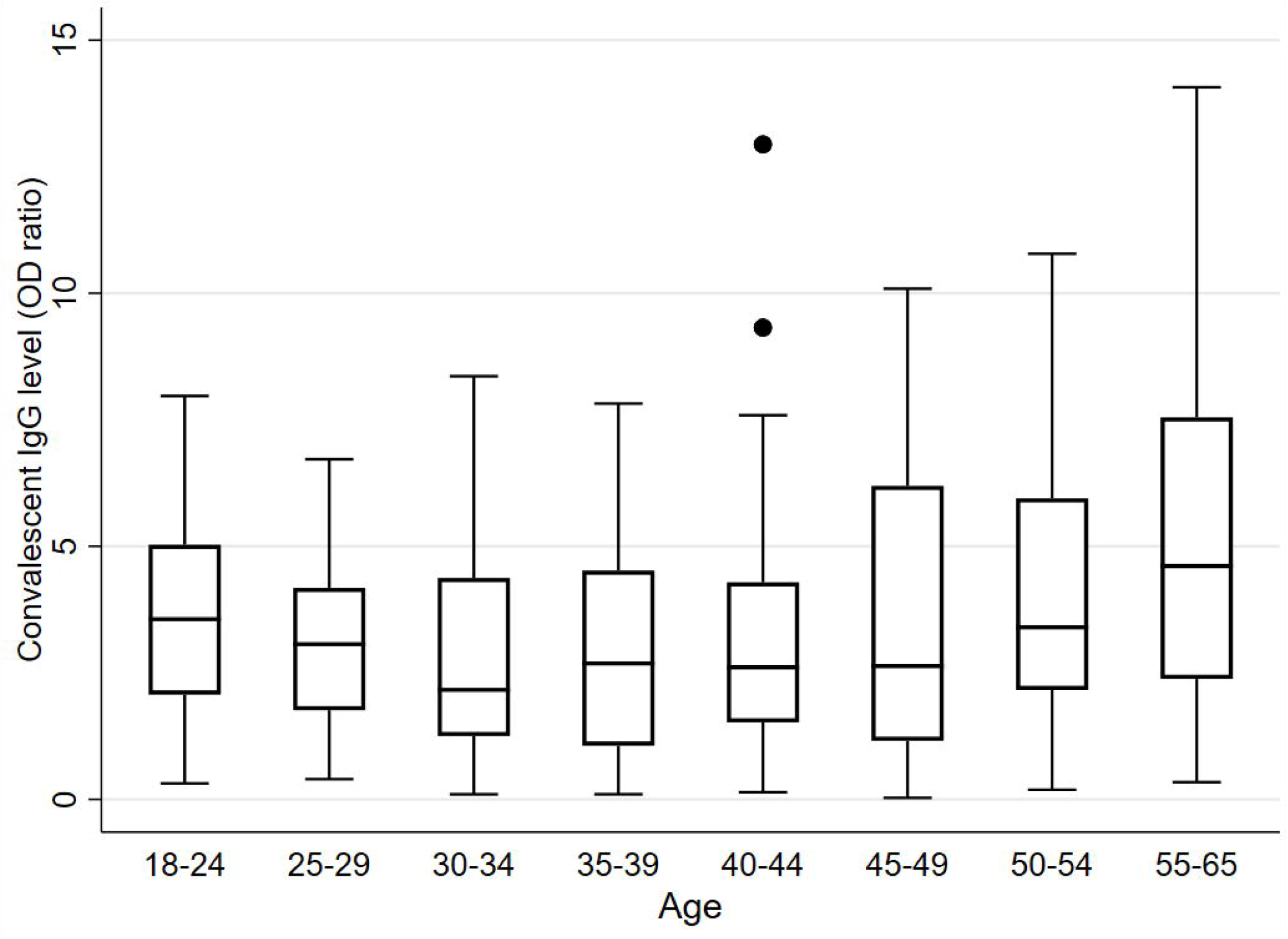
Convalescent SARS CoV-2 IgG by age category. The IgG OD ratio at the convalescent visit by age of the participant.

**Figure 2.**
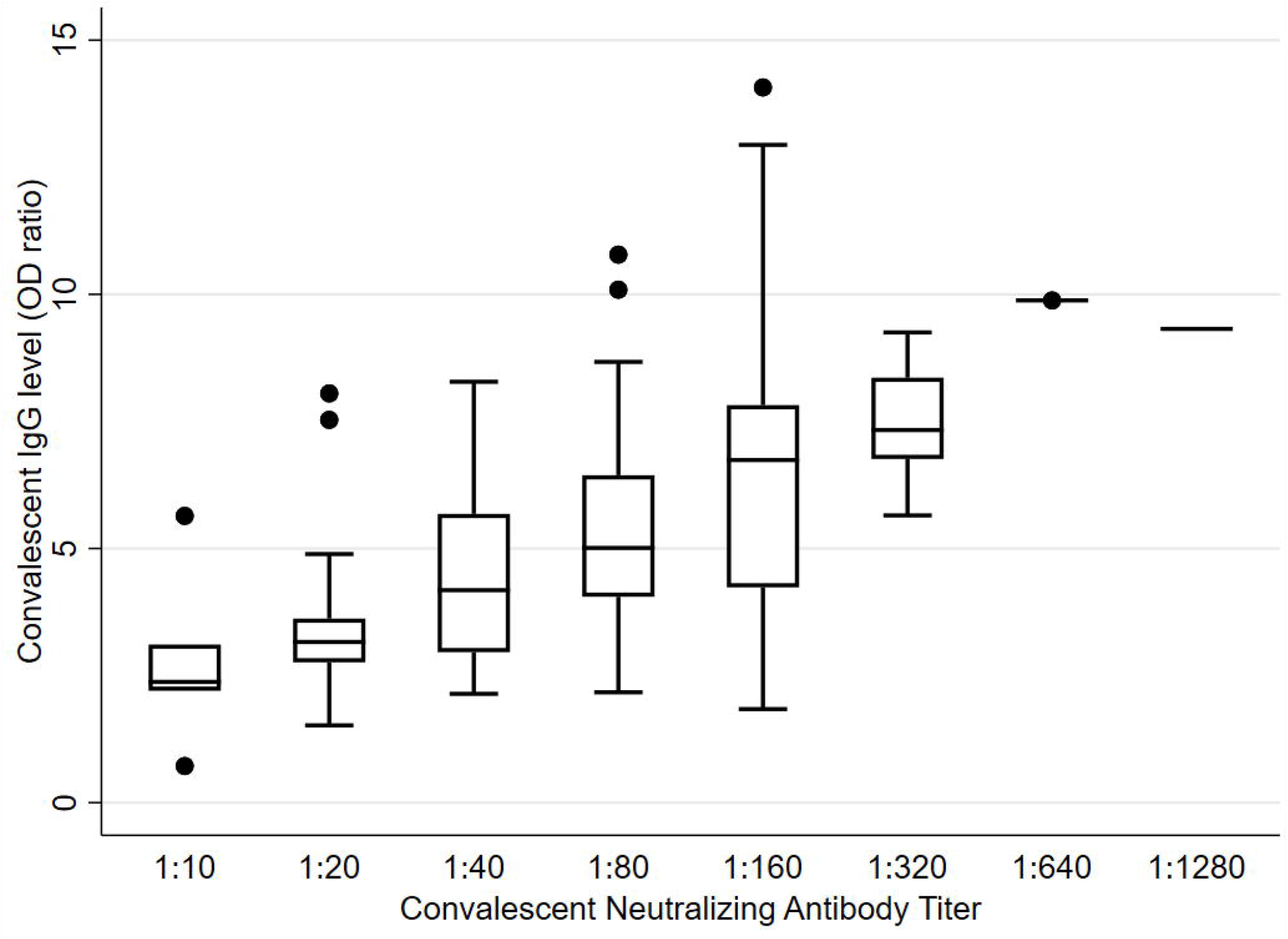
Convalescent SARS CoV-2 Neutralizing antibody and IgG titers. The IgG OD ratio compared to the Neutralizing antibody titer at the convalescent visit.

The results of the multiple linear regression model on 200 patients with IgG results and information from the survey showed that higher IgG antibody titers were associated with age 45 to 60 years (0.8, 95%CI 0.05-1.6, p=0.036), compared to age 30 to 44) and moderate/severe activity restriction (2.1, 95%CI 1.2-3.0, p<0.001), compared to normal activity) (Table 2). A similar analysis carried out on the 120 participants with survey results and nAb titers showed that male gender (2.2, 95% CI 0.9-5.6, p=0.097, compared to female) and a CT-value of 25-30 (4.9, 95%CI 0.9-25.8, p=0.062, compared to CT-value>30) from the diagnostic swab were associated with high nAb titers in the univariate logistic regression model at the 0.1 level (Table 3 and Supplemental Figure 3).

For the 63 participants tested with the nucleocapsid protein ELISA, IgG levels were classified as negative for 24 (38.1%), 11 (17.5%) as weak positive, and 28 (44.4%) as high positive. The correlation between the spike protein and nucleocapsid protein assays was 73.0% (Kappa=0.548) and the nucleocapsid assay and nAbs was 16.7% (Kappa=0.022) (Supplemental Figure 4).

The illness severity score was analyzed separately as it combined multiple variables that were already included in the regression models. Based on the categories outlined above, 32.3% (52/161) were defined as mild, 54.0% (87/161) as moderate, and 13.7% (22/161) as severe illness. Participants with severe disease had median IgG titers of 6.0 (IQR 3.8 to 7.6), compared to 2.5 (IQR 1.4 to 4.1) for those with mild disease and 2.9 (IQR 1.5 to 4.7) for moderate disease (p<0.001). For nAbs, we observed a trend that participants with more severe disease had high titers, but the difference was not statistically significant (OR 1.9, 95%CI 0.9-3.8, p=0.084).

The 35 participants eligible for the follow-up testing had the second blood collection a median of 46 days (IQR 23 to 57) after the convalescent testing. The median IgG OD ratio was 3.7 (IQR 2.9 to 5.9), and 74.3% (26/35) were categorized as having a high positive IgG response. The IgG titers showed a median increase of 0.5 (IQR −0.3 to 1.7) from convalescent to follow-up testing (p<0.001) (Table 1 and Figure 3). The nAbs titers from the six follow-up participants were all ≤ 1:80 (Figure 4).

**Figure 3.**
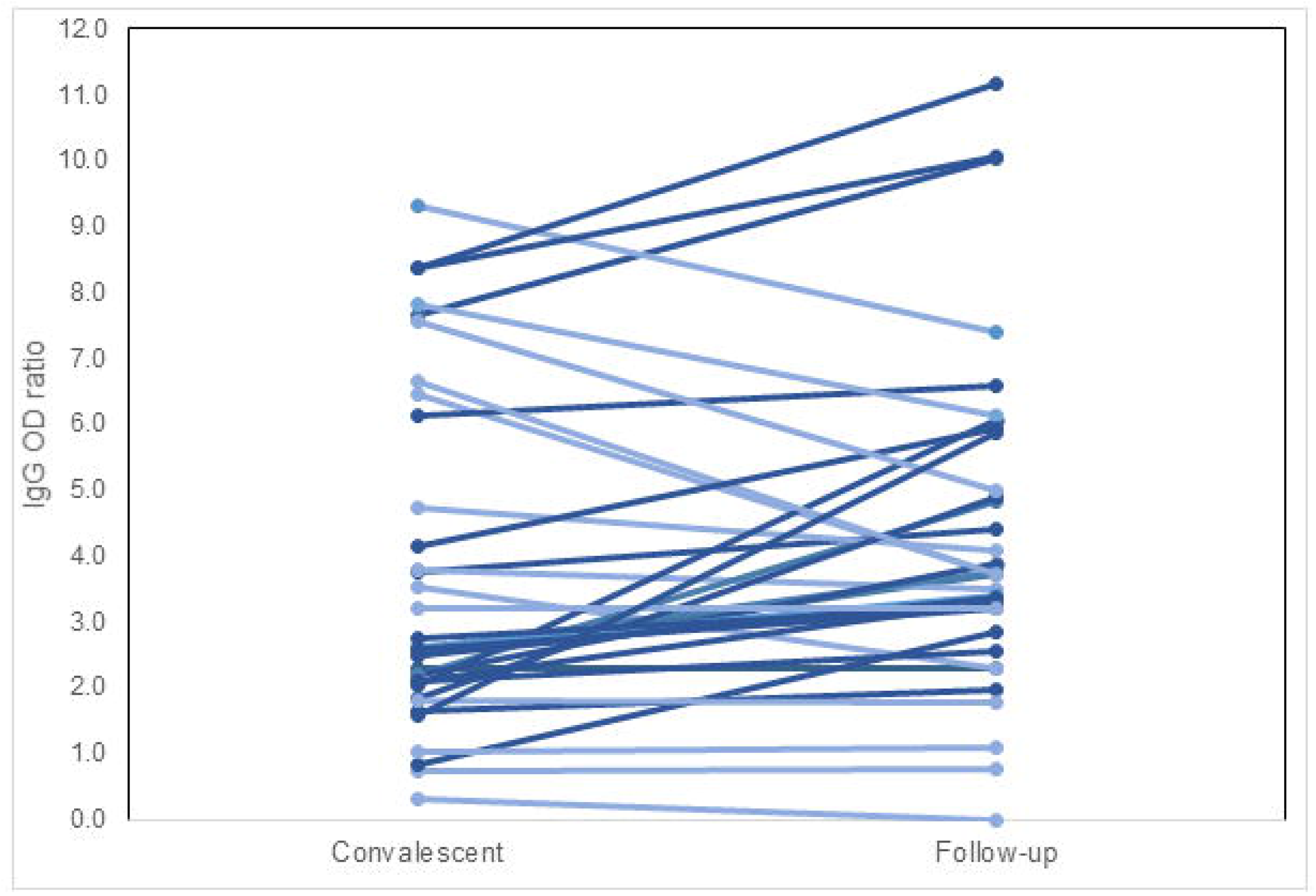
Change in SARS CoV-2 IgG levels from Convalescent to Follow-up testing. The change in IgG OD ratio from the convalescent to the follow-up time points for 35 participants with available data. Dark blue lines indicate increasing OD ratio, and light blue lines indicate the same or decreasing OD ratio.

**Figure 4.**
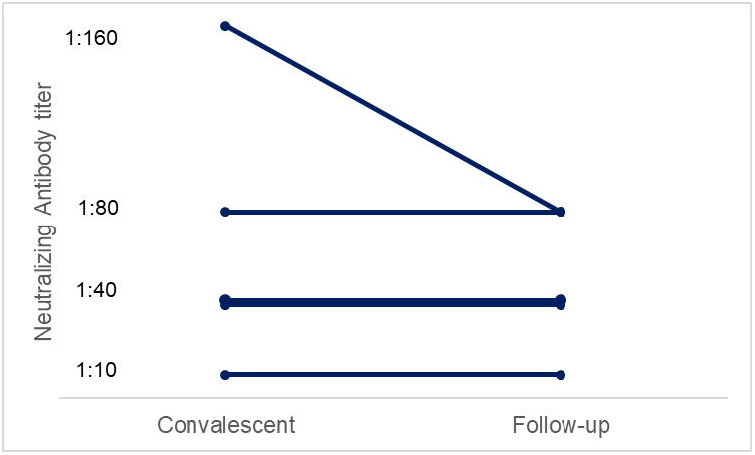
Change in SARS CoV-2 Neutralizing antibody from Convalescent to Follow-up testing. The change in neutralizing antibody titers from the convalescent to the follow-up time points for 5 participants with both timepoints available. There are two participants with 1:40 titers at both Convalescent and Follow-up timepoints.

## Discussion

This study describes the antibody response in a cohort of 281 recovered COVID-19 patients. Our results show that older age (45-65 years) and more severe disease were associated with a stronger IgG response. This response was sustained up to 47 days after the convalescent sample in those who had a follow-up visit. However, there was poor correlation between clinical characteristics and the strength of a neutralizing antibody response. Furthermore, in the few participants that had nAbs follow-up testing, a reduction in the neutralization capacity of the circulating IgG antibodies was observed. The persistence of nAbs over time must be confirmed in future studies with larger cohorts.

We found 15.3% of patients not to have an antibody response at initial testing. This finding is similar to a finding in a study of over 1,300 SARS-CoV-2 patients in New York City that reported IgG seroconversion to the S protein in 89% PCR-positive patients at the time of initial testing from Wajnberg et al. Our observation of the IgG response being correlated with age and disease severity confirms the findings of a prior study by Klein et al, where older age and hospitalization was associated with stronger anti-SARS-CoV-2 antibody response quantified by both IgG levels against the S protein and nAb titers [10]. In this study of 126 plasma donors, male sex and older age were also observed to be associated, however we were only able to deduce an association of nAbs but not IgG levels with male sex.

While our study showed persistence of an IgG response after a month and a half in those with high initial IgG titers, a study of 365,000 adults in England by Ward et al. showed a decline in the prevalence of SARS-CoV-2 antibodies over the course of three months. The decline in IgG positivity was greatest among those who reported no symptoms, a population that was underrepresented in our study [16].

These findings have implications for the selection of convalescent donors of plasma for plasma therapy [13]. Thus, in order to improve the anti-SARS-CoV-2 IgG titers present in the donated plasma, donors should be selected from those with a more severe course of COVID-19 disease and asked to donate plasma as soon as possible after convalescence [10, 17]. Furthermore, these findings might have implications for immunity against repeat infections. However, further data is needed to understand whether antibodies or T cells confer protective immunity to SARS-CoV-2.

At the convalescent study visit, 9% of participants remained positive for SARS-CoV-2 on PCR. However, all CT-values were >30, indicating a low viral load and likely non-viable virus particles. These data are in line with other studies reporting positive PCR results up to 28 days and longer after symptom resolution [3, 18-20]. Surprisingly, persistent viral shedding was not associated with IgG or nAbs response in our analysis, supporting the idea that shed RNA is not an indicator of ongoing infection [3, 8].

This study has several limitations. The criteria for plasma donation restricted participation to healthy people under 60 years of age. Further, the survey responses showed that most participants had relatively mild COVID-19 disease, although asymptomatic participants were underrepresented and very few were hospitalized. These factors indicate that the results may not be generalizable to populations who are older than 60, have severe disease, have comorbidities, or to asymptomatic patients. Most participants only had one serum sample collected, so there is limited data on the change in antibody titers over time. The variability in the nAb response would require evaluation of larger numbers of convalescent patients in order to yield a better understanding of predictive factors.

This study also has several strengths. We present the IgG response in over 200 participants and nAbs in 145 participants. We also collected follow-up data for those participants who had a strong IgG and nAbs response at baseline. The information on illness severity and PCR results from both diagnosis and convalescent time-points provided a fuller clinical picture.

## Conclusion

While older age and disease severity were correlated with stronger IgG responses, the predictive factors for strong neutralizing antibody response in convalescent patients remain unclear.

## Data Availability

De-identified data and analysis code are available upon request to the corresponding author.

## Footnote page

### Funding

This work was supported by the German Federal Ministry of Education and Research (BMBF) program on emergency research funding for COVID-19 (funding number 01KG20152) and internal funds of Heidelberg University Hospital.

### Author contributions

CMD was responsible for the conceptualization and design of the study and supervised the analysis and writing. MG was responsible for the statistical analysis and writing. CMT, PK and TB were responsible for the literature review and supported the conceptualization and design of the study. PK, YH, LR, JS, MJ, NB, and TS were responsible for the survey and patient recruitment. SW and MJ were responsible for the study visits and blood collection. HGK, RB, MSC, BM and AP were responsible for the antibody testing. All authors contributed to data interpretation, critically reviewed the first draft, and approved the final version and agreed to be accountable for the work.

### Ethical Approval

All procedures performed in studies involving human participants were in accordance with the ethical standards of the institutional research committee of Heidelberg University (S686/2018) and with the 1964 Helsinki declaration and its later amendments or comparable ethical standards.

## Acknowledgements

We would like to thank all the participants who contributed to the survey and plasma donation study. We are grateful to Maria Anders-Össwein, Stefanie Wolf, and Ira Pistorius for their dedicated expert technical assistance.

