## Supplemental Figures for "Can we predict antibody responses in SARS-CoV-2? A cohort analysis"

### Supplemental Material

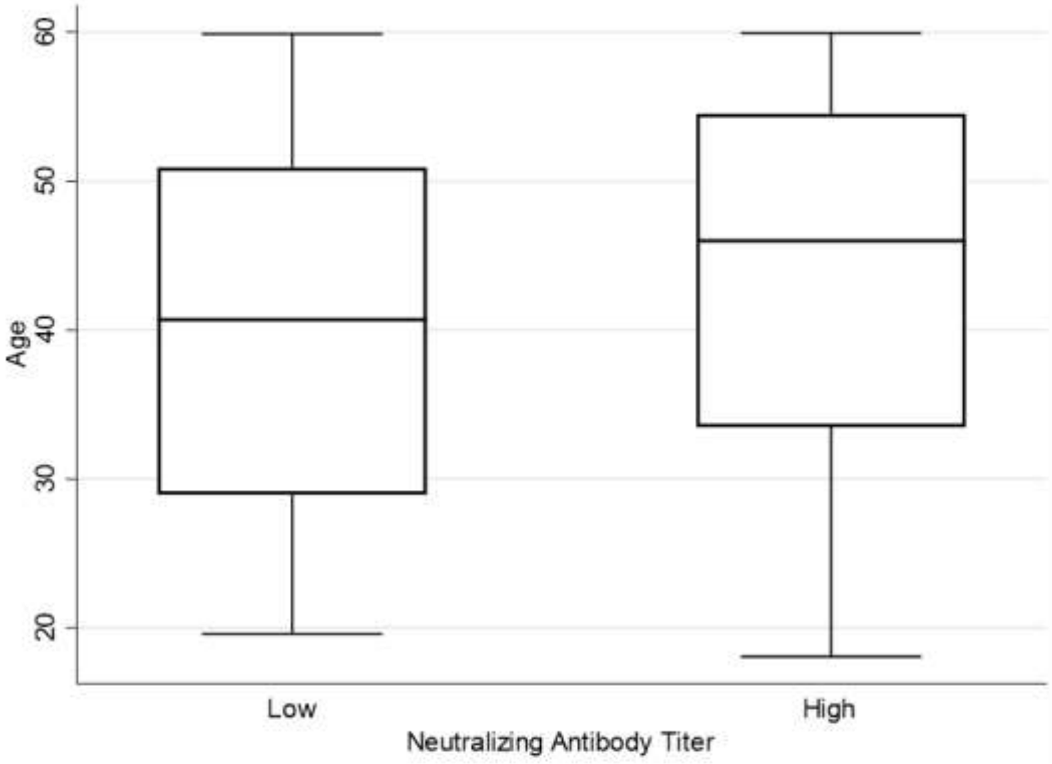


**Supplemental Figure 1. Convalescent SARS CoV-2 Neutralizing AB titers by age**

*The Neutralizing antibody titer at the convalescent visit by age of the participant. Low≤1:80; High>1:80*

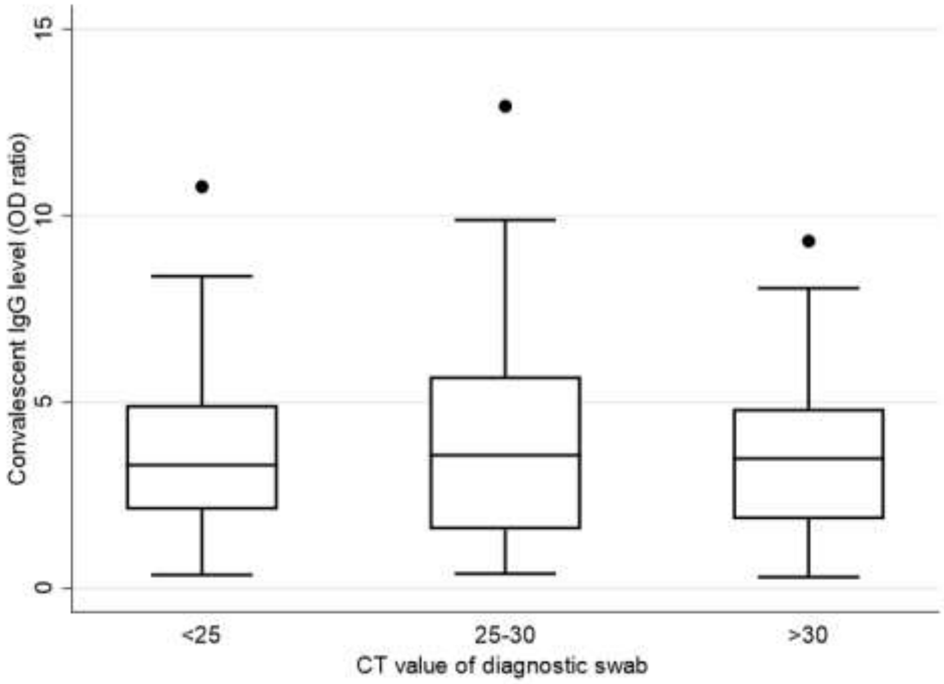


**Supplemental Figure 2. CT value of diagnostic PCR by SARS CoV-2 Convalescent IgG levels**

*The convalescent IgG OD ratios compared to the PCR CT values of the diagnostic test.*

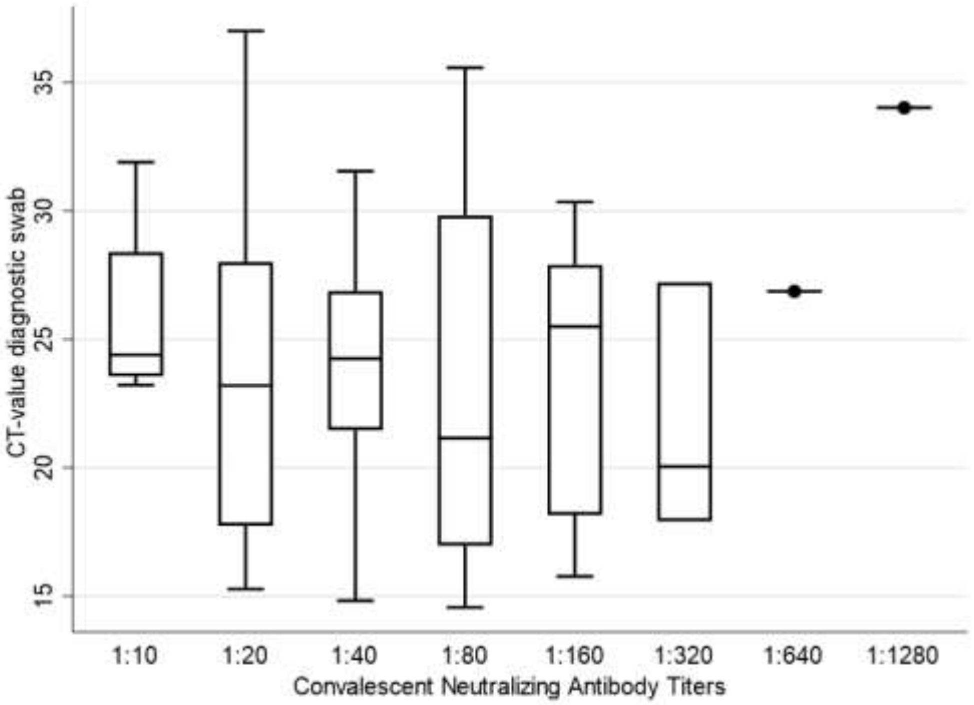


**Supplemental Figure 3. CT value of diagnostic PCR by SARS CoV-2 Neutralizing Antibody titers**

*The convalescent Neutralizing antibody titers compared to the PCR CT values of the diagnostic test.*

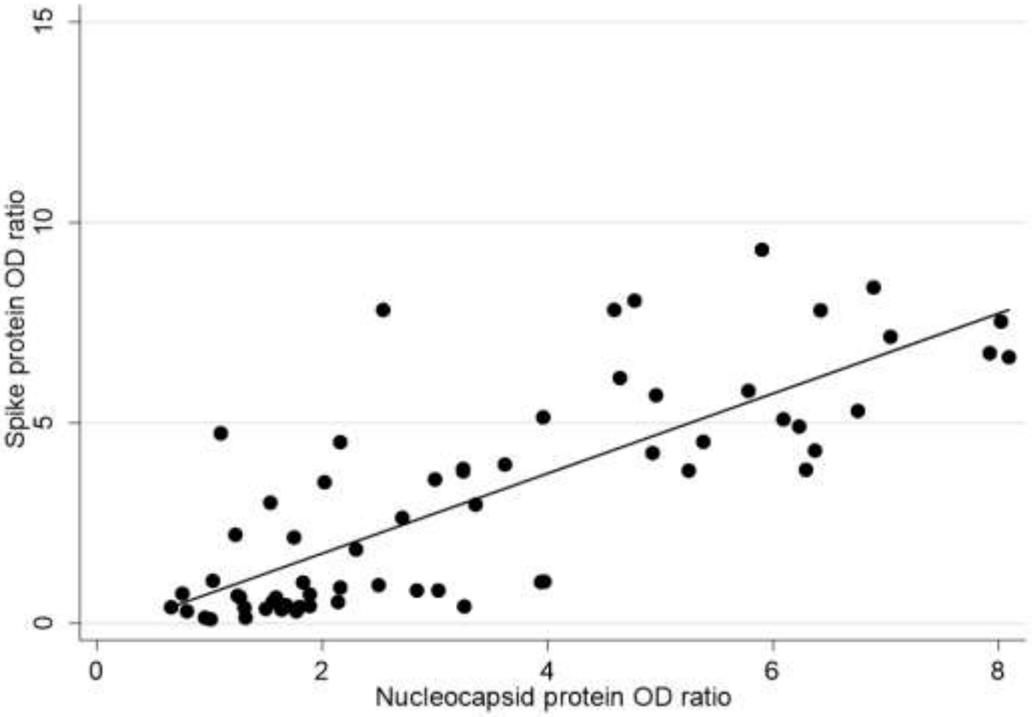


**Supplemental Figure 4. Comparison of SARS-CoV-2 Spike protein and Nucleocapsid protein assays**

*The IgG OD ratios of the spike protein assay (Euroimmune) compared to the nucleocapsid protein assay (Immunodiagnostics) for 63 participants with trend line.*
